# On the assessment of deep-learning based super-resolution in small datasets of human brain MRI scans

**DOI:** 10.64898/2026.02.16.26346392

**Authors:** Dirk W.M. Loeffen, Anne Rijpma, Ronald H.M.A. Bartels, R. Saman Vinke

## Abstract

Deep-learning based super-resolution has shown promise for enhancing the spatial resolution of brain magnetic resonance images, which may help visualize small anatomical structures more clearly. However, when only limited training data are available, it remains uncertain which model assessment method provides the most reliable estimate of out-of-sample performance. In this study, three widely used assessment strategies (*three-way holdout, k-fold cross-validation*, and *nested cross-validation*) were compared for evaluating the performance of such models in small datasets. Across 30 iterations, we randomly selected subsets of 20 T2-weighted images from the 1,113 scans of the Human Connectome Project. Each subset was used to train a model and estimate performance using the three methods. The ground truth error was computed from the remaining images. The assessment error is the difference between the estimated error and the ground truth error. The median assessment errors were 0.11,− 0.13, − 0.32 for *three-way holdout, k-fold cross-validation*, and *nested cross-validation*, respectively, with the cross-validation methods showing considerably smaller dispersions. *Nested cross-validation* selected fewer epochs, indicating more conservative model selection, but required substantially greater computational time, over three times longer than *three-way holdout* and more than twenty times longer than *k-fold cross-validation*. Our findings suggest that *k-fold cross-validation* offers the most favourable balance between accuracy, stability, and computational feasibility in small datasets. Further research is needed to determine how model complexity, dataset size, and the number of cross-validation folds influence assessment accuracy.

## 1. Introduction

### 1.1. Super Resolution for brain MRI

Spatial resolution is generally defined as the finest discernible detail within an image [1]. In neuroimaging, this refers to the ability to distinguish between brain structures within a scan. There is increasing interest in imaging progressively smaller anatomical structures, including brain aneurysms [2], the subthalamic nucleus [3], and early-stage brain tumours [4].

In medical imaging, spatial resolution is sometimes erroneously equated with pixel count [5]. Pixel count merely defines the upper bound of achievable spatial resolution; the actual spatial resolution is determined by the imaging process itself and the highest frequency information captured during acquisition. Examples of methods that allow higher frequency information to be captured are increasing scanning time or improving hardware.

However, these traditional approaches of capturing high-frequency information are not always feasible. Extending the scanning duration is inconvenient for patients and increases vulnerability to motion artefacts. It also places additional pressure on healthcare systems, as it increases workload within radiology departments. Improving hardware is similarly challenging. For example, increasing the magnetic field strength can increase problems such as geometrical distortion [6]. Furthermore, such hardware upgrades may not be financially viable for all hospitals.

Alternatively, spatial resolution can be improved by using software-based methods, particularly deep-learned networks. The breakthrough of the convolutional neural network (CNN) for image analysis occurred in 2012 [7]. This led to widespread adoption in medical application, such as medical image classification and later medical image super-resolution. The first application of a CNN for super-resolving brain MRI scans produced impressive results [8], initiating a new research line that ultimately resulted in an FDA-cleared super-resolution software package [9]. For this reason, we opt to employ deep-learning-based methods for super-resolution in this study.

For deep-learned super-resolution networks, it is essential to estimate performance on new, unseen images. Numerous approaches exist for such assessment. In this paper, we evaluate three widely used methods: *three-way holdout, k-fold cross-validation* and *nested cross-validation. Three-way holdout* is generally recommended for model selection in deep learning when datasets are sufficiently large [10]. When datasets are small, however, this approach may be suboptimal. Since a smaller dataset necessitates a smaller test set, and from the central limit theorem it follows that the average of a small test set is more likely to deviate from the true population average. More recent papers also experimentally confirmed this theoretical concern [11]. Background information on all three assessment strategies is provided in Appendix B. Their implementation is described in the sections 2.5, 2.6 and 2.7. In clinical practice, small MRI datasets are particularly common, as patient recruitment is challenging and data collection places additional strain on radiology departments. This issue becomes even more pronounced when addressing highly specific research questions. Consequently, we aim to investigate these assessment methods specifically in the context of small MRI datasets. The focus of this paper is therefore on hospital patient populations, as MR super-resolution is primarily applicable in this setting.

### 1.2. Objective

Although there are indications that *nested crossvalidation* provides a more accurate estimate of the out-of-sample performance of deep-learned models compared with *three-way holdout* or *k-fold crossvalidation*, this has not yet been demonstrated in deep-learning-based super-resolution [11]. There-fore, we aim to determine whether *nested crossvalidation* offers a superior assessment of the out-of-sample performance of deep-learned superresolution networks compared with *three-way hold-out* and *k-fold cross-validation*. We hypothesize that *nested cross-validation* provides a more reliable estimate of model performance then both *k-fold cross-validation* and *three-way holdout* when evaluating deep-learned super-resolution on small datasets of brain magnetic resonance images.

## 2. Methods

The experimental protocol was preregistered at the Open Science Framework (OSF). The manuscript was prepared following the Consensus-based Recommendations for Machine-learning-based Science (REFORMS) [12], relevant excerpts have been documented in Appendix A. All code developed for this study, together with instructions for use, is available at Github or alternatively at the OSF project of this study.

### 2.1. Data

Open-source data from the human connectome project (HCP) were used in this study [13]. The HCP cohort consists of 1113 young healthy adults aged 22–35, from whom 1113 structural 3 Tesla (3T) magnetic resonance scans were obtained. The initial target size was 1200 scans; the reason this number was not reached has not been reported. The outcome variable of interest, the T2-weighted (T2w) 3T scans, were acquired between October 2015 and August 2022 at the Washington University using a modified Siemens 3T Skyra scanner. The corresponding MR acquisition parameters are listed in Table 1 [13].

**Table 1:**
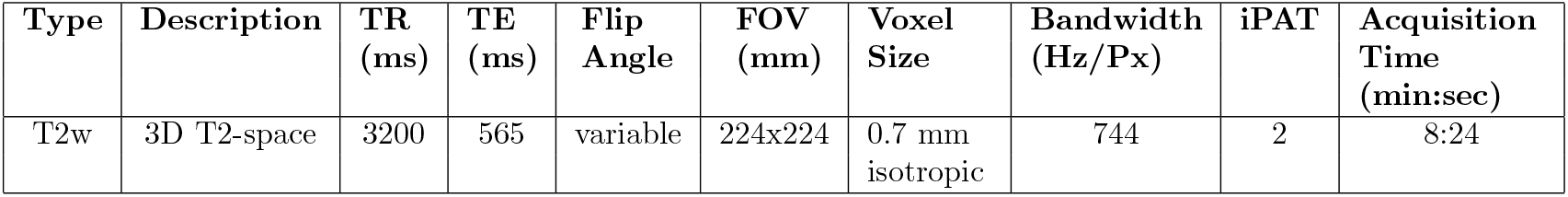
Magnetic resonance parameters of the T2-weighted 3T scans from the human connectome project (HCP) [13].

| Type | Description | TR<br>(ms) | TE<br>(ms) | Flip<br>Angle | FOV<br>(mm) | Voxel<br>Size | Bandwidth<br>(Hz/Px) | iPAT | Acquisition<br>Time<br>(min:sec) |
| --- | --- | --- | --- | --- | --- | --- | --- | --- | --- |
| T2w | 3D T2-space | 3200 | 565 | variable | 224x224 | 0.7 mm<br>isotropic | 744 | 2 | 8:24 |

The sampling frame comprised young adult sibships of average size three to four, with many sibships containing twins. Sibship including individuals with severe neurodevelopmental disorders, documented neuropsychiatric disorders or neurological disorders were excluded. No additional exclusions were applied for the purpose of the current study. Although the presence of twins technically violates the assumption that all data points are independent and identically distributed, we argue that this does not influence our experiments. Namely, when sub-sampling 20 subjects from the full dataset, at most 20 co-twins could appear in the remaining dataset (*n* = 1093), which constitutes a negligible proportion of the external dataset.

While the HCP sampling frame is not a perfect representation of the clinical population described in the introduction, we consider the dataset appropriate for our purposes. The MRI scans exhibit comparable anatomical complexity to clinical scans, making the performance of different assessment methods comparable. For imaging, we used the T2w processed images registered to native T1w space. All scans were anonymized by blurring the face and the ears. Since we did not want this blur to influence the training process, we used the skull-stripped images available within the HCP dataset^1^.

### 2.2 Preprocessing

During preprocessing we did not detect any impossible or corrupt samples. This section summarizes the HCP preprocessing steps and describes all additional preprocessing applied in the present study. Full details of the HCP structural preprocessing pipeline can be found in [14]. The T2w images used underwent the PreFreeSurfer and FreeSurfer pipeline. In short, the gradient distortion was corrected, the T2w images were aligned to the T1w images, the brain was extracted, readout distortion was corrected, and bias-field correction was applied.

Additional preprocessing for this study was implemented using functions from the monai package [15]. DICOM folders were loaded using *LoadImage*, images were reoriented to RAS orientation using *Orientation*, and intensities between the 0.1 and 99.9 percentiles were rescaled to the range 0 to 255 using *ScaleIntensityRangePercentiles* to produce the high-resolution images. The images were then blurred using *GaussianSmooth* with a sigma of 1 voxel, and downsampled by a factor of 2 in each spatial dimension using *Spacing* to produce low-resolution images. The low-resolution images were subsequently resampled to match the high-resolution image shape using *ResampleToMatch*. Both high- and low-resolution images were saved as NIfTI files using *SaveImage*. No dependencies between training and testing data were introduced during preprocessing.

### 2.3. Data Loading

Data loading for training also utilized functions from the *monai* package. Each low-resolution image and its corresponding high-resolution image were loaded as a pair. Using *RandomSpatialCropSamplesd*, 200 random paired image patches were sampled from each low-/high-resolution pair. Sampling patches increases the number of training samples and reduced the risk of overfitting. Low-resolution patches served as model input, and high-resolution patches served as ground truth.

For validation and testing, full low-resolution and high-resolution image pairs were loaded without patch sampling. The full low-resolution images were fed to the models as input, and the high-resolution images served as ground truth.

### 2.4. Training Details

We used the ReCNN model proposed by [8] for super-resolution, owing to the simplicity of its training and the extensive evaluation by the authors. During training, the input consisted of 64 × 64 × 64 LR patches, and the output consisted of corresponding 64 × 64 × 64 super-resolved patches. For validation, internal testing and external testing, full LR images were used as input, producing full super-resolved outputs. Model outputs were compared with HR patches or images using a mean squared error (MSE) loss function, the MSE is defined in Appendix C.

To limit the training duration, we set the number of layers to 5, used a filter size of 3 × 3 × 3 and used 32 filters per layer. Weights and biases were initialized using the default initializer of the *Conv2D* layer, which samples from a uniform distribution between 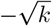 and 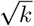 where *k* is the number of kernel elements. The Adam optimizer was used with a learning rate of 1^−3^. All training procedures were implemented in the *monai* python package.

Models were trained on a dedicated computing cluster running *Ubuntu 22.04.2 LTS*. The cluster included an *AMD Ryzen Threadripper PRO 3995 WX CPU* with 64 cores, 512 GB of random-access memory (RAM) and a 1000 GB hard drive for data storage. All code was executed using Python 3.9. Parallelised computations were performed on three *NVidia GeForce RTX 3090 GPUs*. Multi-GPU training was managed using *PyTorch Lightning* ^2^.

### 2.5. Three-way holdout

As the baseline assessment method, we used three-way holdout, which is standard in the field. For each iteration, the 20 selected subjects were divided into training, validation and test sets using the ratios 0.6, 0.2, and 0.2, respectively. The model was trained on the training set and validated each iteration on the validation set using the MSE. Training continued until the validation MSE failed to improved for 15 iterations. Increases in MSE of less than 0.01 were considered negligible. Once the stopping criterion was reached, the model was retrained for the same number of iterations using both the training and validation sets, allowing the model to benefit from the additional data. The test-set MSE was used as the estimate for the out-of-sample performance. After this estimate was obtained, the model was retrained on all 20 selected subjects with the same number of epochs and the true out-of-sample performance was given by evaluating the final model on the out-of-assessment set. The final measurement was the difference between the performance estimate and the true out-of-sample performance.

### 2.6. k-fold cross-validation

For *k*-fold cross-validation, the 20 selected subjects were partitioned into *k* = 5 groups. For each of the five iterations, a model was trained on 4 folds and validated on the remaining fold. Training proceeded according to the same stopping criterion described in Section 2.5. The performance estimate was the mean of the five final validation MSE values. The median number of training iterations was then used to retrain the model on all 20 subjects to obtain the true out-of-sample performance on the out-of-assessment dataset. The final measure was the difference between the performance estimate and the true out-of-sample performance.

### 2.7. Nested cross-validation

For nested cross-validation, the 20 selected subjects were divided into *n* = 5 groups. Each group served once as the test set. For a given test fold, the remaining 4 groups were used to perform an inner 4-fold split, such that each group served once as a validation set and the remaining three formed the training set. This procedure resulted in 4 × 5 trained models. Training for each model followed the stopping criterion described in Section 2.5. The median number of epochs from the inner loop was used to retrain a model on the combined training and testing groups, and the performance was assessed on the corresponding the test set. This produced 5 performance assessments, one per test group. The final performance estimate was the mean of these 5 assessments. The median of the five median epoch counts was then used to retrain the final model on all 20 subjects, and true out-of-sample performance was assessed on the out-of-assessment subjects. The final measure was the difference between the performance estimate and the true out-of-sample performance measure.

### 2.8. Analysis

Every model evaluation method was repeated 30 times. For each iteration, we obtained both a performance estimate and a true out-of-sample performance. Their difference constituted the *model assessment error*. We hypothesized that *k*-fold cross validation would yield a biased estimate and that three-way holdout would have higher variance relative to nested cross-validation. We visualized the distributions of model assessment error and reported the mean, median and first and third quartiles. Contrary to protocol, 95% highest density intervals were not reported, owing to an insufficient number of observations.

We further compared the mean squared model assessment error, as this metric penalizes both the bias and the variance and is therefore well-suited for comparing assessment methods [10]. We plotted the distributions of mean squared model assessment error, reported the mean and standard deviation, and reported the median with the first and third quartiles. Again, 95% highest density intervals were not reported due to insufficient observations.

Because the experiment could, in principle, be repeated an arbitrary number of times, null hypothesis significance testing was not a meaningful analytic strategy. Instead, we focused on the substantive differences between the assessment methods and present these differences using descriptive statistics.

In addition to the above analysis as already described in the preregistered analysis, we also report the number of training epochs and the total assessment time required for each evaluation method. Although these measures were not included in the original protocol, we consider them informative for readers, as they provide insight into the practical feasibility of the assessment strategies. The number of epochs reflects how long each model was trained before meeting its stopping criterion, thereby offering an indirect indication of model convergence behaviour across the three assessment methods. It is important to understand that convergence will happen earlier in small datasets, since there is less to learn. The total assessment time reflects the computational cost of each method, which is particularly relevant when evaluating methods such as nested cross-validation that require multiple model trainings. Reporting these two measures therefore gives a more complete overview of the trade-offs between statistical robustness and computational burden for each assessment method.

## 3. Results

### 3.1. Assessment Error

The distributions of the assessment error are shown in Figure 1. The assessment error was defined as the difference between the estimated out-of-sample performance subtracted by the true out-of-sample performance. Consequently, negative values indicate underestimations of the assessment error. For *three-way holdout*, the mean assessment error was −0.02±1.18, the median was 0.11 (0.69−−0.65), and the range was [−2.67, 2.37]. For *k-fold cross-validation*, the mean assessment error was −0.10 ± 0.69, the median was −0.13 (0.17 − −0.48), with a range of [−1.62, 1.65]. For *nested cross-validation*, the mean assessment error was −0.43 ± 0.67, the median was −0.32 (−0.09 − −0.81), and the range was [−2.53, 1.13].

**Figure 1.**
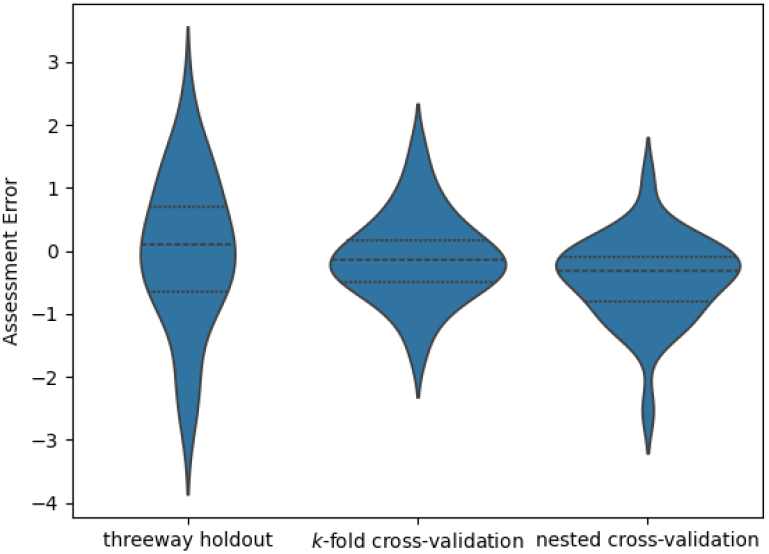
Violin plots showing the distribution of the assessment error of the three different assessment methods. The error is calculated as internal error subtracted by external error. In consequence, negative values are underestimations of the performance.

### 3.2. Absolute Assessment Error

The distributions of the absolute assessment error are displayed in Figure 2. *Three-way hold-out* produced a mean absolute assessment error of 0.91 ± 0.72, a median of 0.69 (1.42 − 0.33), and a range of [0.03, 2.67]. For *k-fold cross-validation*, the mean absolute assessment error was 0.53± 0.44, the median was 0.40 (0.68 − 0.17), and a range of [0.06, 1.65]. For *nested cross-validation*, the mean absolute assessment error was 0.58± 0.55, the median was 0.37 0.90 − 0.21, and the range was [0.03, 2.53].

**Figure 2.**
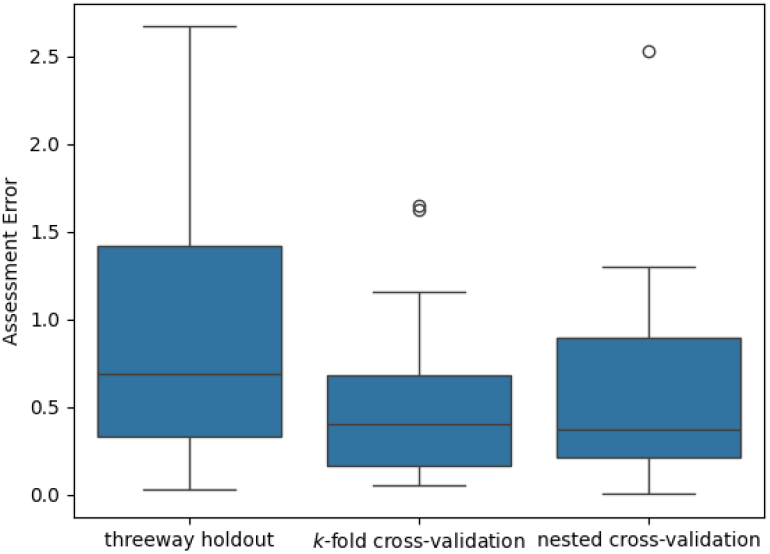
Box plots showing the distribution of the absolute assessment error of the three different assessment methods.

**Figure 3.**
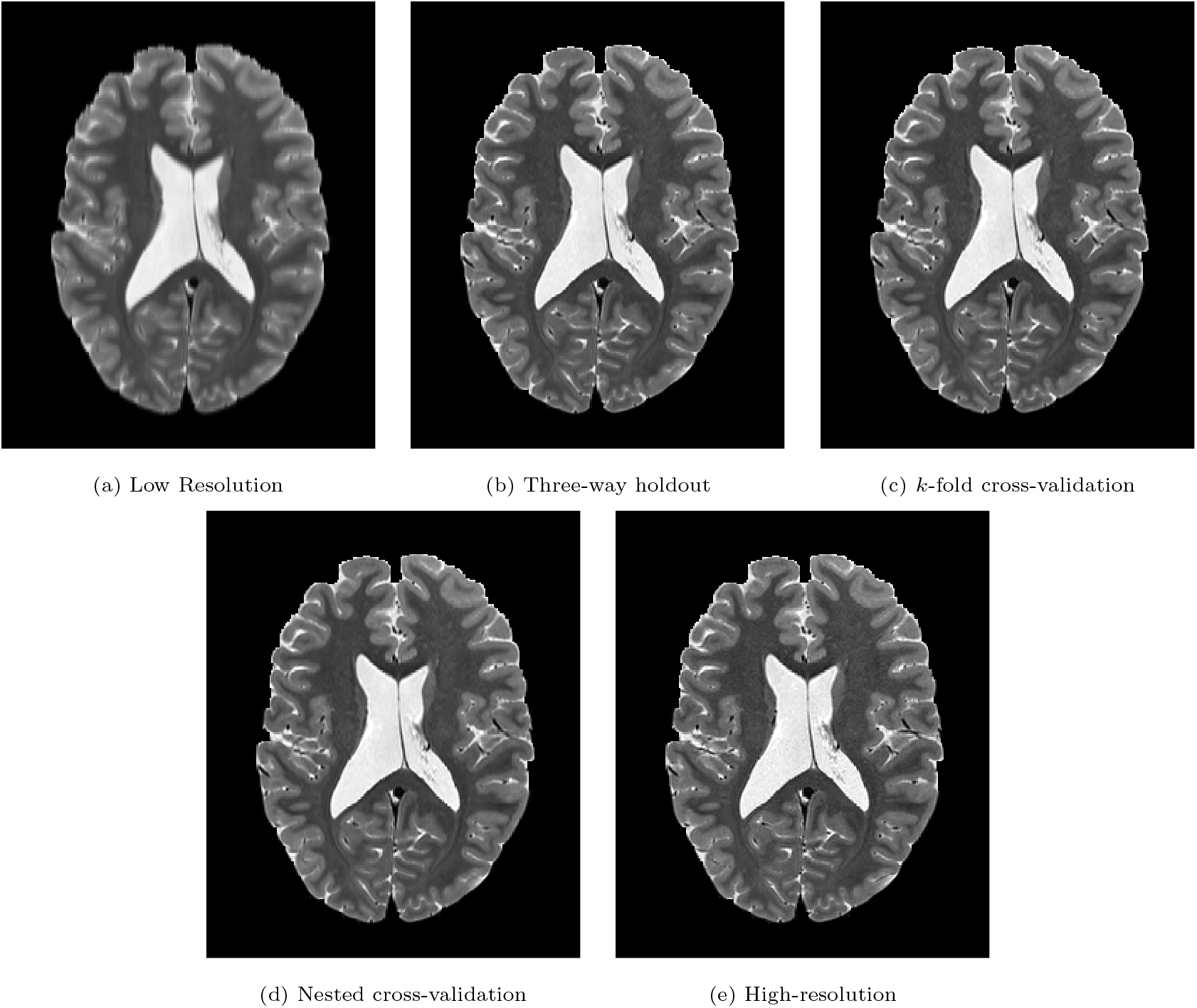
Example of a low-resolution input, the results from the three different assessment methods and the corresponding high-resolution image.

### 3.3. Number of Epochs

For *three-way holdout*, the median number of epochs was 40.5 (48 − 35.25), and the range was [22, 60]. For *k-fold cross-validation*, the median number of epochs was 39 (42 − 36.25), with a range of [28, 45]. For *nested cross-validation*, the median number of epochs was 33.5 (36 − 30.25), and the range was [20, 59].

### 3.4. Assessment Time

For *three-way holdout*, the median total assessment time was 1.20 hours (1.45 − 1.10), with a range of [0.67, 1.81]. For *k-fold cross-validation*, the median was 8.43 hours (10.13 − 7.77), with a range of [6.49, 11.69]. For *nested cross-validation*, the median was 27.21 hours (32.69 − 23.55), and the range was [18.46, 40.65].

## 4. Discussion

In this study, we show that *k-fold cross-validation* provides a more accurate estimate of model per-formance than *nested cross-validation* when using small datasets. One possible explanation is that *nested cross-validation* relies on fewer data points for hyperparameter selection and therefore behaves more conservatively, as reflected in the smaller number of training epochs selected. In contrast, the *three-way holdout* approach displayed substantially greater dispersion in its estimates when compared with the other two methods. This indicates that individual instances of *three-way holdout* can lead to much larger under- or overestimations of performance.

We originally hypothesized that *nested crossvalidation* would offer the most accurate performance estimates for deep-learning based brain MRI super-resolution, on the basis that *k-fold crossvalidation* may introduce data leakage and potential overfitting to the validation folds, and that *three-way holdout* relies solely on a single split of the data. However, our findings demonstrate that *k-fold cross-validation* produced more accurate performance estimates *nested cross-validation* in the present small-dataset setting. These results do not imply that *k-fold cross-validation* is universally preferable to either *nested cross-validation* or *three-way holdout*.

Firstly, *k-fold cross-validation* may perform better only under the specific conditions examined here. We trained a shallow model on MRI data, which are typically complex and high-dimensional. It is plausible that the models did not overfit substantially during *k-fold cross-validation*, whereas deeper architectures with more trainable parameters might overfit more readily. Future work should investigate at which model complexity *k-fold cross-validation* begins to yield biased performance estimates.

Secondly, it must be acknowledged that both *nested cross-validation* and *k-fold cross-validation* involve a bias-variance trade-off and are not generally unbiased estimators of out-of-sample performance. Only in the special case where *k* is the total number of samples, i.e. leave-one-out cross-validation, that these are both unbiased. For *nested cross-validation*, such an approach would be computationally infeasible, as it would require training a prohibitively large number of models. Increasing *k* for *k-fold cross-validation*, would lead to a larger variance in performance estimates. Researchers must therefore make an informed decision based on the relative importance of bias and variance in their specific application.

Another consideration is that the optimal number of epochs was determined using fewer samples in *nested cross-validation* than in *k-fold crossvalidation*. As shown in Section 3.3, *nested crossvalidation* selected notably fewer epochs on average, suggesting a more conservative training regime. This may be beneficial if minimizing overfitting is the primary goal, which would make *nested crossvalidation* attractive in some settings. Therefore, if reducing overfitting is the primary goal, *nested cross-validation* might be the better choice.

Despite its conservative behaviour, the substantially longer assessment time required for *nested cross-validation* makes it impractical for routine use. As reported in Section 3.4 the median assessment time was more than three times higher than that of *k-fold cross-validation* and more than 22 times higher than that of *three-way holdout*. With models with more trainable parameters, the total computation time would grow rapidly, potentially extending to several months, making the method infeasible for many research projects.

The results further show that *three-way holdout* far greater dispersion in its estimates than either of the cross-validation methods in small datasets. This is particularly concerning since *three-way hold-out* yields a single point estimate without an associated uncertainty interval, making this variability invisible to the researcher. Based on these findings, we recommend that researchers adopt methods that provide a distribution or interval estimate rather than relying solely on a single-split assessment.

This study raises several opportunities for further research. As noted above, it is important to examine whether the current findings extend to deeper models with more trainable parameters. Additionally, it would be valuable to determine the minimum sample size at which *three-way holdout* becomes reliable relative to the cross-validation approaches. Finally, an open methodological question is how the choice of the number of folds in both the *k-fold* and *nested cross-validation* affects the accuracy of performance estimate.

## 5. Conclusion

In this study, we compared three commonly used methods for assessing the performance of deeplearning super-resolution models in small datasets. Our results show that *k-fold cross-validation* provided the most accurate and stable performance estimates on average. Although *three-way hold-out* yielded competitive point estimates in some instances, its substantially greater dispersion makes it less reliable, particularly because it provides no measure of uncertainty. *Nested cross-validation* produced more conservative performance estimates and offers conceptual advantages for model selection; however, its markedly higher computational cost limits its practicality in many research settings.

Taken together, our findings suggest that *k-fold cross-validation* represents an effective balance between accuracy, stability, and computational feasibility when evaluating deep-learning-based super-resolution models in small datasets of brain MRI. Nevertheless, *nested cross-validation* remains a valuable alternative when conservative estimates are desired or when the risk of overfitting is of particular concern. Researchers should therefore choose an assessment method based on their specific priorities; whether this is accuracy, uncertainty estimation, or computational efficiency.

## Data Availability

All data produced are available online at https://doi.org/10.17605/OSF.IO/J6QXD and at https://github.com/dwml/SuperResAssess

https://www.humanconnectome.org/study/hcp-young-adult

## 6. Declaration of generative AI and AI-assisted technologies in the manuscript preparation process

During the preparation of this work the authors used Microsoft Copilot to edit the final version of the manuscript for legibility. After using this Microsoft Copilot, the authors reviewed and edited the content as needed and take full responsibility for the content of the published article. The manuscript as prepared prior to the use of Microsoft Copilot can be found here

## Appendix A. REFORMS reporting guide-line

### Appendix A.1. Module 1: Study Goals

1a. Population or distribution about which the scientific claim is made.

Section 1.1: “The focus of … in this setting

1b. Motivation for choosing this population or distribution (1a.).

Section 1.1: “Consequently, we aim … small MRI datasets”

1c. Motivation for the use of ML methods in the study.

Section 1.1: “The first application … super-resolution software package.”

#### Appendix A.2. Module 2: Computational Reproducibility

2a. Dataset used for training and evaluating the model along with link or DOI to uniquely identify the dataset.

Section 2.1, footnote 1.

2b. Code used to train and evaluate the model and produce the results reported in the paper along with link or DOI to uniquely identify the version of the code used.

Section 2.4, footnote 2.

2c. Description of the computing infrastructure used.

Section 2.4: “Models were trained … using PyTorch Lightning.”, and footnote 2.

2d. README file which contains instructions for generating the results using the provided dataset and code.

At our GitHub page.

2e. Reproduction script to produce all results reported in the paper1.

Since the first step of the reproduction is to download the HCP files, for which the user needs to set credentials, we cannot provide a full reproduction script. The README on our GitHub page provides information on how to reproduce our experiment. Additionally, the GitHub page provides information on how to reproduce the images.

#### Appendix A.3. Module 3: Data Quality

3a. Source(s) of data, separately for the training and evaluation datasets (if applicable), along with the time when the dataset(s) are collected, the source and process of ground-truth annotations, and other data documentation.

Section 2.1: “Open-source data from … in Table 1 [13].”

3b. Distribution or set from which the dataset is sampled (i.e., the sampling frame).

Section 2.1: “Open-source data from … in Table 1 [13].”

3c. Justification for why the dataset is useful for the modeling task at hand.

Section 2.1: “While the HCP … assessment methods comparable.”

3d. The definition of the outcome variable of the model along with descriptive statistics, if applicable.

Section 2.1: “Open-source data from … in Table 1 [13].”

3e. Number of samples in the dataset.

Section 2.1: “Open-source data from … in Table 1 [13].”

3f. Percentage of missing data, split by class for a categorical outcome variable.

Section 2.1: “Open-source data from … in Table 1 [13].”

3g. Justification for why the distribution or set from which the dataset is drawn (3b.) is representative of the one about which the scientific claim is being made (1a.).

Section 2.1: “The sampling frame … the external dataset

#### Appendix A.4. Module 4: Data Preprocessing

4a. Identification of whether any samples are excluded with a rationale for why they are excluded. Section 2.1: “No additional exclusions … the current study”

4b. How impossible or corrupt samples are dealt with.

Section 2.2: “During preprocessing, we … or corrupt samples.”

4c. All transformations of the dataset from its raw form (3a.) to the form used in the model, for instance, treatment of missing data and normalization.

Section 3.2: “DICOM folders were … files using SaveImage.”

#### Appendix A.5. Module 5: Modelling

5a. Detailed descriptions of all models trained, including:

Section 2.4

5b. Justification for the choice of model types implemented.

Section 2.4: “We used the … by the authors.” 5c. Method for evaluating the model(s) reported in the paper, including details of train-test splits or cross-validation folds.

Sections 2.5, 2.6, and 2.7.

5d. Method for selecting the model(s) reported in the paper.

Sections 2.5, 2.6, and 2.7.

5e. For the model(s) reported in the paper, specify details about the hyperparameter tuning:

Sections 2.5, 2.6, and 2.7.

5f. Justification that model comparisons are against appropriate baselines.

Section 2.5: “As the baseline … in the field.”

#### Appendix A.6. Module 6: Data Leakage

6a. Justification that pre-processing (Section 4) and modeling (Section 5) steps only use information from the training dataset (and not the test dataset). Section 2.2: “No dependencies between … introduced during preprocessing.”

6b. Methods to address dependencies or duplicates between the training and test datasets (e.g., different samples from the same patients are kept in the same dataset partition).

Section 2.1: “The sampling frame … the external dataset.”

6c. Justification that each feature or input used in the model is legitimate for the task at hand and does not lead to leakage.

Since in this paper only raw MRI images are used this is not applicable

#### Appendix A.7. Module 7: Metrics and Uncertainty

7a. All metrics used to assess and compare model performance (e.g., accuracy, AUROC etc.). Justify that the metric used to select the final model is suitable for the task.

Section 2.8: “For each iteration, … to insufficient observations”

7b. Uncertainty estimates (e.g., confidence intervals, standard deviations), and details of how these are calculated.

Section 3

7c. Justification for the choice of statistical tests (if used) and a check for the assumptions of the statistical test.

Section 2.8: “Because the experiment … using descriptive statistics.”

#### Appendix A.8. Module 8: Generalizability and Limitations

8a. Evidence of external validity.

Since no claims about future performance are made this is not applicable

8b. Contexts in which the authors do not expect the study’s findings to hold.

Section 5: “Firstly, k-fold cross-validation … a single-split assessment.”

## Appendix B. Assessment Methods

In this section, we discuss the mathematical setting for the *nested cross-validation* assessment method more extensively. Since we expect most readers to be aware of the assessment methods this has been moved to the appendix. For ease of notation, we use the mean as the centrality measure in all formulas, but this could be changed for the median without a difference. In the current study, the training of the super-resolution model will be performed on a dataset *I* that holds *n* pairs of low and high-resolution images, *lr*_*i*_ and *hr*_*i*_ respectively:

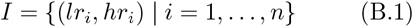

The model is denoted *s* and its parameters are denoted *θ*. We would like to find a model with the optimal parameters and they are denoted 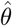. To get 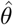, we minimize some loss function *L* with respect to the parameters *θ*, formulated as:

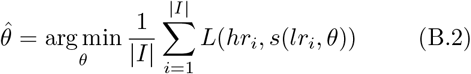

When *s* has sufficient capacity then *ŝ* can theoretically fit the dataset perfectly. When the model fits our current dataset well, this does not mean the model will predict well when used on images that are not in our dataset. One way of estimating this generalization error is by using *k-fold cross-validation*.

### Appendix B.1. k-fold cross validation

To estimate the generalization error of our optimal model *ŝ* using *k-fold cross-validation*, we use:

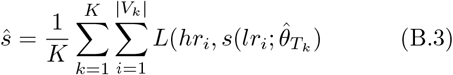

Here 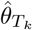 are the optimal parameters given *T*_*k*_ and 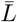 is the estimated generalization error. A depiction of the *k-fold cross-validation* is shown in Fig B.4. One problem is still that in principle we could train the model indefinitely. For *k-fold cross-validation* we decide to stop based on the generalization error estimate; when this does not decrease, we stop training. This is not the neatest stopping criterion, since it introduces a dependency between all *V*_*k*_ and 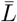. A method that does not introduce this dependence is the three-way holdout.

#### Appendix B.2. Three-way holdout

When using three-way holdout the dataset is split in training set *T*, validation set *V*, and test set *A*, this is depicted in Fig B.5. As noted in the last section, it is unknown how many training iterations we should use. The number of iterations, or epochs, is an example of a hyperparameter. In the current work it is the only hyperparameter we tune, so we can denote this *ϵ*. Now we can find the optimal number of epochs 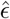, using:

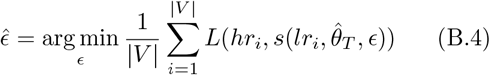

With the optimal parameters *thêta*_*T*_ found equivalently as before using the training set *T*. Now the independence between the parameters and the number of epochs has been maintained and we can estimate the generalization error, using:

**Figure B.4:**
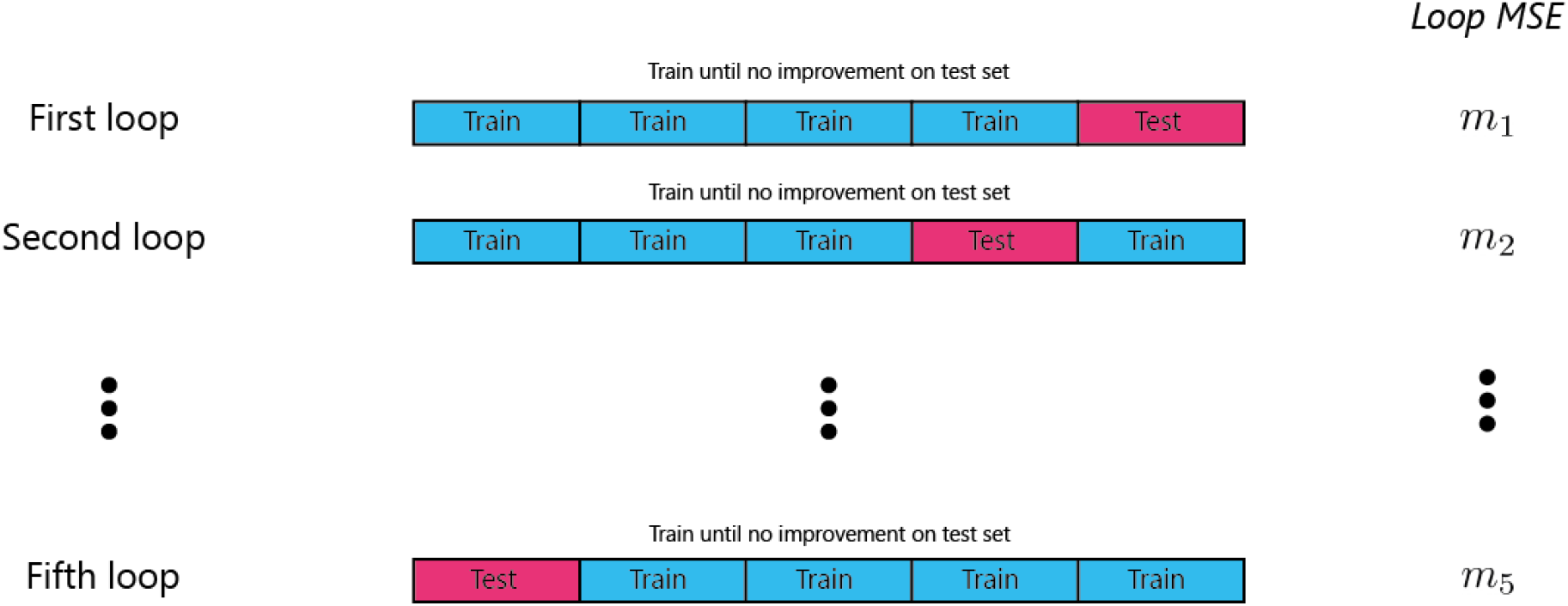
Depiction of how data is used when using the k-fold cross-validation assessment method with *k* = 5. *MSE: mean squared error*

**Figure B.5:**
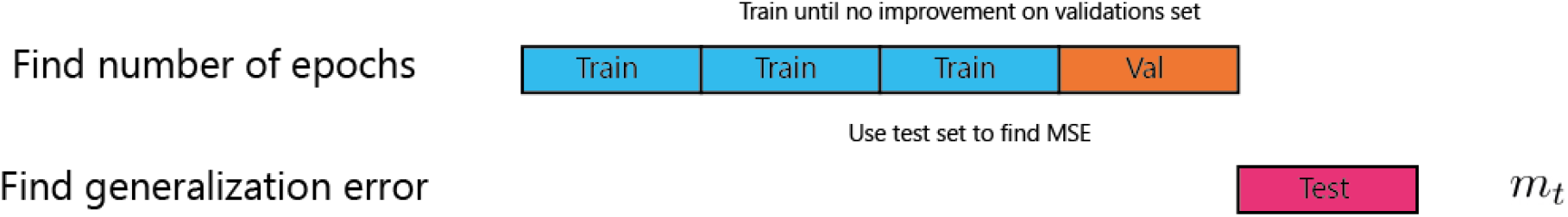
Depiction of how data is used when using the three-way holdout assessment method. *MSE: mean squared error*

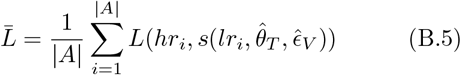

This is a good estimator of the true generalization error, given that the test set is sufficiently large [10]. If it is not sufficiently large, the probability that it does not reflect the true generalization error becomes increasingly high with decreasing size. In that case, the *nested cross-validation* might be a better method.

#### Appendix B.3. Nested cross-validation

To better estimate the true generalization error, Stone came up with the cross-validatory assessment of the cross-validatory choice of the model, this is what we call the *nested cross-validation* [16]. It is a double *k-fold cross-validation*, where we first make folds for the test set, for the cross-validatory assessment, then we make folds for the validation set, for the cross-validatory choice of the model. This whole procedure is depicted in Fig B.6. Nested cross-validation is sometimes called *k* × *l* cross-validation, hence we will use the subscripts *k* and *l*. The test set will be *A*_*k*_ for the *k*th outer loop, and the training and validation set will be *T*_*l*_ and *V*_*l*_ for the *l*th inner loop. For the assessment of the generalization error, we can now write:

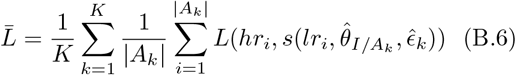

In this formula, *K* is the total number of folds and 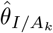are the parameters found by retraining a model on the dataset without the respective test set, with the mean number of epochs found in iteration *k*:

**Figure B.6:**
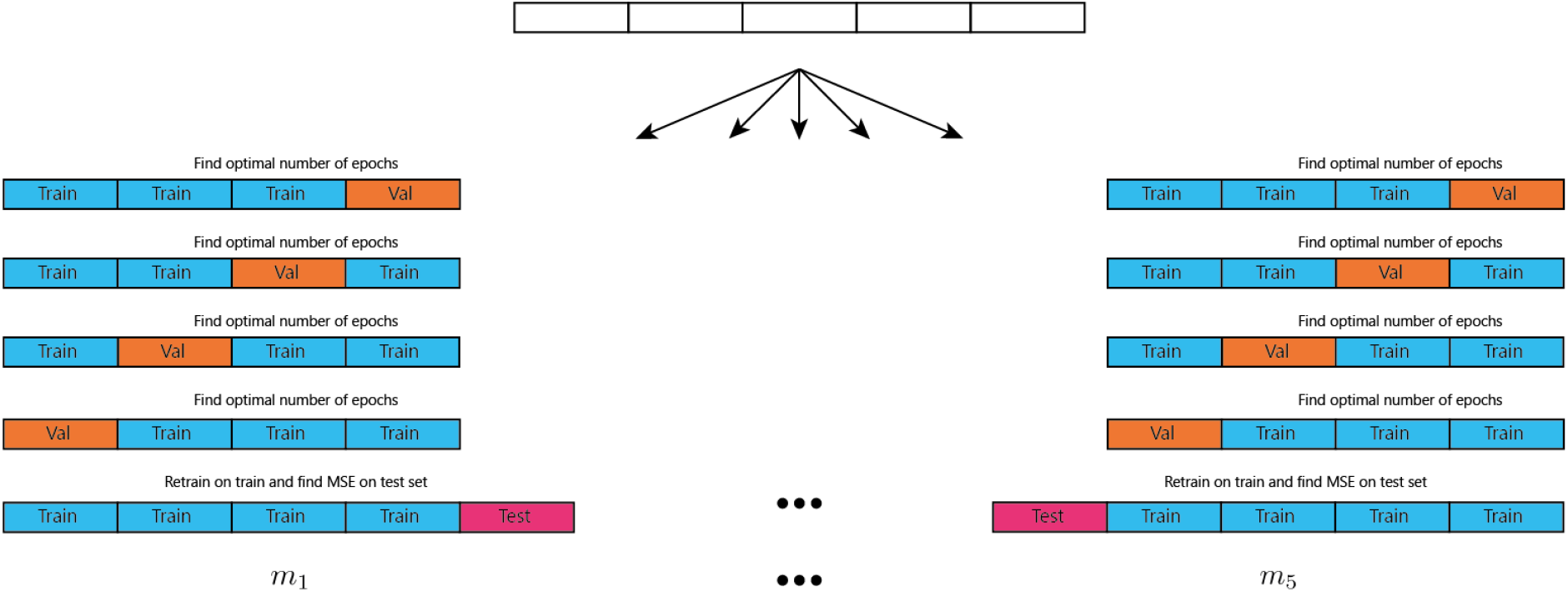
Depiction of how data is used when using nested cross-validation in this case *K* = 5 and *L* = 4. *MSE: mean squared error*

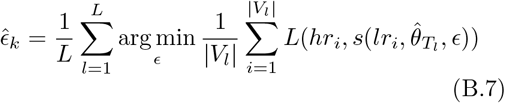

## Appendix C. Mean Squared Error

In the current study the mean squared error (MSE) is defined as:

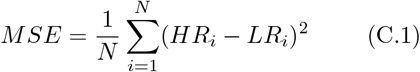

*HR* is the high-resolution image, *LR* is the low-resolution image and *N* is the number of image pairs in the set that the MSE is calculated on where *i* is the index of this pair.

## Footnotes

1 The exact set of images used can be found at our OSF project: https://osf.io/j6qxd/files/vm2f6.

2 All versioning details can be found at our OSF project: https://osf.io/j6qxd/files/qb2dh

